# A machine learning based predictive model for the diagnosis of sepsis

**DOI:** 10.1101/2022.04.29.22274361

**Authors:** Juan A. Delgado Sanchis, François Signol, Juan-Carlos Perez-Cortes, Eva M. García-López, Salvador Mena-Mollá, Nieves Carbonell-Monleón, María Rodríguez-Gimillo, José Luis García-Giménez

## Abstract

The early recognition and treatment of sepsis is essential to increase the probability of survival of the patient. Sepsis is a complex and heterogeneous syndrome influenced by the site of infection, causative microorganisms, acute organ dysfunctions and co-morbidities. So, early diagnosis is a challenge in which complex physiologic, metabolic, biochemical markers and clinical signs must be evaluated simultaneously for a reliable and early identification of sepsis. In this paper, a list of relevant variables involved in sepsis diagnosis is provided. Furthermore, to help answering the question of whether a patient is suffering from sepsis when entering the emergency room, a model based on the machine learning gradient boosting algorithm is proposed. Using three histones H2B, H3, H4 and the activated protein C together with other variables a gradient boosting classifier was trained and evaluated with cross validation. Results show that the model can achieve up to a 97% mean per-class accuracy.

## 1. Introduction

The reliable and early identification of sepsis is often complicated by its own syndromic nature and not always the availability of microorganismal positive test which may contribute to delays in treatment. When a patient is admitted to an emergency room, it is interesting to have a predictive model fed with variables available at admission time and capable of helping in the early diagnosis of sepsis. Ideally, knowing how to distinguish between sepsis and any other confounding pathological entity. Detection of sepsis at admission time and its treatment is essential to reduce mortality (Evans et al. 2021) and several studies that implement artificial intelligence (AI) techniques are associated with a low mortality rate. This fact indicates that AI models could decrease the time to detect sepsis significantly, providing a benefit to the health of the patient (Schinkel et al. 2019).

The objectives of this study are: 1) to identify variables that could be relevant in sepsis diagnosis using an ensemble feature selection algorithm. A particular interest is put on the three histones H2B, H3, H4 and the activated protein C delivered by the kit HistShock® 2) to build a predictive model that answers the question whether a patient is suffering sepsis. The model could be used as a support decision system for early diagnosis.

The rest of this paper is organized as follows. Section 2 describes the dataset, the pre-processing phase, the feature selection and classification technique employed. Sections 3 and 4 list the feature selection and model results with a discussion about the dataset size and robustness of the results; Finally, section 5 draws conclusions and perspectives about this work.

## 2. Material and methods

Data was obtained from the clinical data reports of each participating patient admitted at the Intensive Care Unit (ICU) of the Hospital Universitario Clínico de Valencia (HCUV) after signing informed consent. The study was approved by the Ethical Committee of the HCUV with registry number 2019/051. Patient’s data consisting of about 220 variables from 140 patients was provided in pseudo. The participants in this study form three groups: severe sepsis cases (24 individuals), septic shock (99) and critically ill patients at ICU without suspicion of a septic process, which were set as controls (17). Both cases and controls were admitted in the ICU. Patients with sepsis were admitted via the emergency room.

### 2.1. Dataset

According to the previous criteria for patient selection, a dataset was constructed with the contribution of 220 variables grouped into different types: demographic, clinical, hospitalization and treatment. The information gathered make it possible to study the diagnosis and the evolution of sepsis in the days following admission to an ICU. This raw dataset was pre-processed with the steps described in section 2.2 and then introduced into a machine learning algorithm to help in the sepsis early diagnosis.

The clinical variables included a novel set of variables extracted for each patient by intensivists to improve the detection of sepsis using its proprietary test HistShock®. These variables are histone H2B concentration (ng/mL), histone H3 concentration (ng/mL), histone H4 concentration (ng/mL), and activated protein C (ng/mL).

### 2.2. Pre-processing phase

In this phase, variables provided by intensivists have been pre-processed to build a high-quality dataset in order to generate a machine learning based predictive model of sepsis.

#### 2.2.1. Text normalization

The text normalization steps carried out in this work consisted of capitalize all text, remove accentuation, and manually review categories. These steps were necessary to correct inconsistencies and data entry errors. In this process, a set of useless categories have been discarded.

#### 2.2.2. No histone detection

In this dataset, the histone concentration variables (ng/mL) present low values “0.01”, “0.1”, and “1” which means no histone detection. These outliers were normalized and set to 0.0.

#### 2.2.3. Quality criterion of categorical variables

From a data analysis point of view, it is important that the categories included in the study have sufficient representation. That is, a sufficiently high number of patients in each category. Otherwise, by chance, a category may be explanatory without being able to guarantee that it is really the case or that it is due to the limited size of the database (lack of patients of this category).

A reasonable criterion is to set the minimum size of a category at half the number of patients in the less populated class. In our case, the 17 ICU patients without sepsis corresponded to the less populated class. Consequently, a minimum threshold of 8 patients was established as the minimum frequency for a category to be included in this study. Furthermore, in order not to lose information, it was proposed to group together the categories that are not sufficiently populated (under threshold) in a special category called “OTHER”.

#### 2.2.4. Filtering for missing values

Variables with missing data can be imputed. However, to minimize the artefacts that this imputation could generate, it is reasonable to discard variables exhibiting a high rate of missing values, a situation in which imputation could not be reliable. A 30% missing values threshold is set, meaning that a variable with more than 42 patients without valid values is discarded.

After filtering variables with high rate of missing values, continuous variables are imputed with an average of the values for each variable. In the case of categorical variables, the mode is the imputation method. The underlying idea is to consider a patient with an unknown value as if was an average person, imputing this variable with the mean value of the population.

#### 2.2.5. Converting categorical variables to numeric

The method used is called one hot encoding. A categorical variable of N categories is converted in N boolean variables. A value of 1 is set when the patient belongs to the category, 0 if not. When a patient belongs to several categories as for example when the sepsis is originated from several microorganisms, only the categories (e.g., microorganisms) associated to the patient are set to 1.

The categorical variables of this type are: reason for admission, focus of infection, microorganisms and blood cultures.

#### 2.2.6. Clean dataset

Finally, once removed the variables that were acquired after the diagnosis, applied the categorical quality control and filtered out variables with too many missing values, a clean dataset of 122 sepsis patients, 17 ICU control patients and 56 variables is obtained. Table 1 shows the 56 variables and Table 2 describes the dataset classes distribution.

**Table 1:** List of the 56 variables included in the study.

| Demographic data | Comorbidities and history | HistShock | Constant at admission | Admission analytics Plasma levels | Microbiology |
| --- | --- | --- | --- | --- | --- |
| Age | Diabetes mellitus | Histone H2B | Systolic blood pressure | Albumin | Bacteremia |
| Gender | Dyslipidemia | Histone H3 | Mean arterial pressure | Calcium |  |
| Weight | Tobacco | Histone H4 | Heart rate | Cholesterol |  |
| Height | Alcoholism | Activated protein C | Breathing frequency | Triglyceride |  |
|  | Chronic ischemic heart disease |  |  | Bilirubin |  |
|  | Charlson comorbidity index |  |  | Lactate |  |
|  | Peripheral artery disease |  |  | Phosphorus |  |
|  | Brain disease |  |  | Magnesium |  |
|  | Chronic obstructive pulmonary disease |  |  | Zinc |  |
|  | Chronic liver disease |  |  | Urea |  |
|  | Arterial hypertension |  |  | Creatinine |  |
|  | Corticoids treatment |  |  | Sodium |  |
|  | Immunosuppressants treatment |  |  | Potassium |  |
|  | Hospital admission in the previous 3 months |  |  | Bilirubin |  |
|  | Diagnosis of solid tumor |  |  | Ferritin |  |
|  | Antibiotic use in the 2 weeks prior to ICU admission |  |  | Partial pressure of carbon dioxide |  |
|  |  |  |  | Po2/Fio2 ratio |  |
|  |  |  |  | Reactive Protein C concentration |  |
|  |  |  |  | Procalcitonin |  |
|  |  |  |  | Leukocytes |  |
|  |  |  |  | Polymorphonuclear leukocyte |  |
|  |  |  |  | Hemoglobin |  |
|  |  |  |  | Platelets |  |
|  |  |  |  | Activated partial thromboplastin time |  |
|  |  |  |  | Ph |  |
|  |  |  |  | Chlorine |  |
|  |  |  |  | Glucose |  |

**Table 2:**
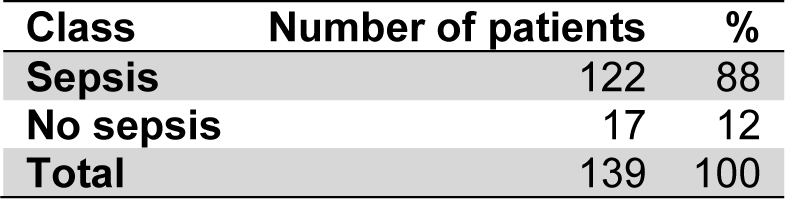
Classes distribution of the dataset

### 2.3. Identify relevant variables with ensemble feature selection

An identification of the relevant variables involved in the sepsis diagnosis at admission time was carried out on the data presented in Table 1 with a feature selection algorithm developed by ITI^1^ based on the ensemble paradigm. This algorithm consists of pooling the opinion of several weak feature selectors to increase the quality and stability of the selection. In this algorithm, a weak selector is a Sequential Feature Selector (SFS) (Pudil, Novovičová, and Kittler 1994) applied to a random train/test stratified split of the data (where proportion of cases and controls is the same than in the original dataset). SFS uses internally a k-neighbors estimator trained with train split and evaluated with test split. It combines variables in a greedy manner and look for the set of them that best predicts the sepsis diagnosis. It results in a ranking of the 56 variables by order of its relevance with the sepsis diagnosis.

### 2.4. Predictive model

The classification of patients with or without sepsis at admission time was done using the Gradient Boosting (GB) machine learning algorithm (Chen and Guestrin 2016) implemented on scikit-learn (Pedregosa et al. 2011) on the dataset obtained in previous section 2.2.4.

GB builds a sequence of decision trees where the errors made in one tree are used to weight the importance of the samples for the next tree. This ensemble method aggregates the selection of characteristics of each tree in a collective decision, which allows the variables to be ordered by their importance in the classification.

### 2.5. Evaluation metrics

The classification quality was evaluated by using a set of measures obtained by comparison of the groundtruth and the predicted class for each patient. The metrics were the following (Zaki and Meira Jr 2014):

- Sensitivity (SEN): also known as True Positive Rate is the ratio of positive events (sepsis/case) that are correctly predicted among all the positive events.
- Specificity (SPC): also called True Negative Rate is the ratio of negative events (no-sepsis/control) which are correctly predicted among all the negative events.
- Balanced Accuracy Score (BACC): also called mean per-class accuracy is the mean accuracy obtained on each class (see Equation 1). The implementation used is the one proposed in [3, 4] which leads a better model generalization measure with imbalanced datasets. This is the case here with 17 non-septic against 123 septic patients.
- Area Under ROC Curve (AUC): The area under the ROC curve, abbreviated AUC, is a measure about the probability of a random positive test case will be ranked higher than a random negative test instance [6].

The gradient boosting classifier was evaluated using both BACC and AUC performance measures. Each learning set is created by taking all the samples except one, the test set being the sample left out. This method is known as leave-one-out cross-validation.

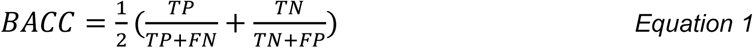

## 3. Results

### 3.1. Relevant variables in sepsis diagnosis

The 56 variables have been ordered by relevance in the diagnosis of sepsis using an ensemble feature selection. This ensemble method splits the dataset into train and test partitions. For each split, train partition is used to build a model and to look for the combination of variables that best predict the test partition. Each appearance of a variable is a vote in its favor and increase its relevance.

It is possible to calculate a threshold above which it is unlikely to obtain this number of votes by luck. This is equivalent to compute the p-value (probability) of gathering a certain number of votes by chance. With a p-value inferior to 0.05, the quantity of votes must be superior to 130. Table 3 shows that three of the six indicators currently used to diagnose sepsis appear in the first positions of the relevant variables list. Focusing on the 4 variables provided by the HistShock test (see section 2.1) circulating histone H2B is clearly considered as relevant with 214 votes while circulating histones H3 and H4 received 109 and 92 votes respectively. H3 and H4 stay below the 130 threshold but received more votes than heart rate, lactate levels at admission and mean arterial pressure which are variables currently used to diagnose sepsis. It suggests that circulating histones H3 and H4 should be further studied.

**Table 3:** Variables ordered by an ensemble voting scheme. 1000 sequential feature selections were calculated leading to a maximum of 1000 votes that can be reached by a variable. The variables with “*” are those used for the diagnosis at emergency room. Variables in bold are the histones measured by the HistShock kit. The first 6 variables exhibit a p-value inferior to 0.05 by obtaining more than 130 votes.

| Variable | Votes |
| --- | --- |
| Systolic blood pressure * | 731 |
| Reactive Protein C levels | 362 |
| Procalcitonin levels * | 333 |
| Bacteremia | 331 |
| **Histone H2B** | 214 |
| Breathing frequency * | 197 |
| Partial pressure of carbon dioxide | 113 |
| **Histone H3** | 109 |
| Patient treated with immunosuppressants | 108 |
| Albumin levels | 103 |
| Antibiotic use in the 2 weeks prior to ICU admission | 102 |
| Heart rate * | 99 |
| **Histone H4** | 92 |
| Chronic ischaemic heart disease | 81 |
| Lactate levels at admission * | 73 |
| Patient treated with corticosteroids | 70 |
| Activated partial thromboplastin time | 66 |
| Hospital admission in the previous 3 months | 62 |
| Charlson Co-morbidity Index | 57 |
| Peripheral artery disease | 55 |
| Sodium levels | 50 |
| Mean arterial pressure * | 46 |
| Bilirubin | 44 |
| Total cholesterol levels | 44 |
| Total bilirubin levels | 44 |
| Chlorine levels | 40 |
| ... | ... |
| **Activated protein C** | 5 |

### 3.2. Sepsis diagnosis modelling

A predictive model helping to diagnose sepsis has been built from the 56 variables. It could be used as a support decision system by consulting the model with the analysis values of a patient as input data.

The gradient boosting used in this experiment in a leave-one-out cross-validation experiment obtained a mean per-class accuracy of 97%. Figure 1 illustrates the sensitivity and specificity curve, showing that it is possible to completely detect all the cases of sepsis (true positives) if we can allow for a 20% of false positive rate.

**Figure 1:**
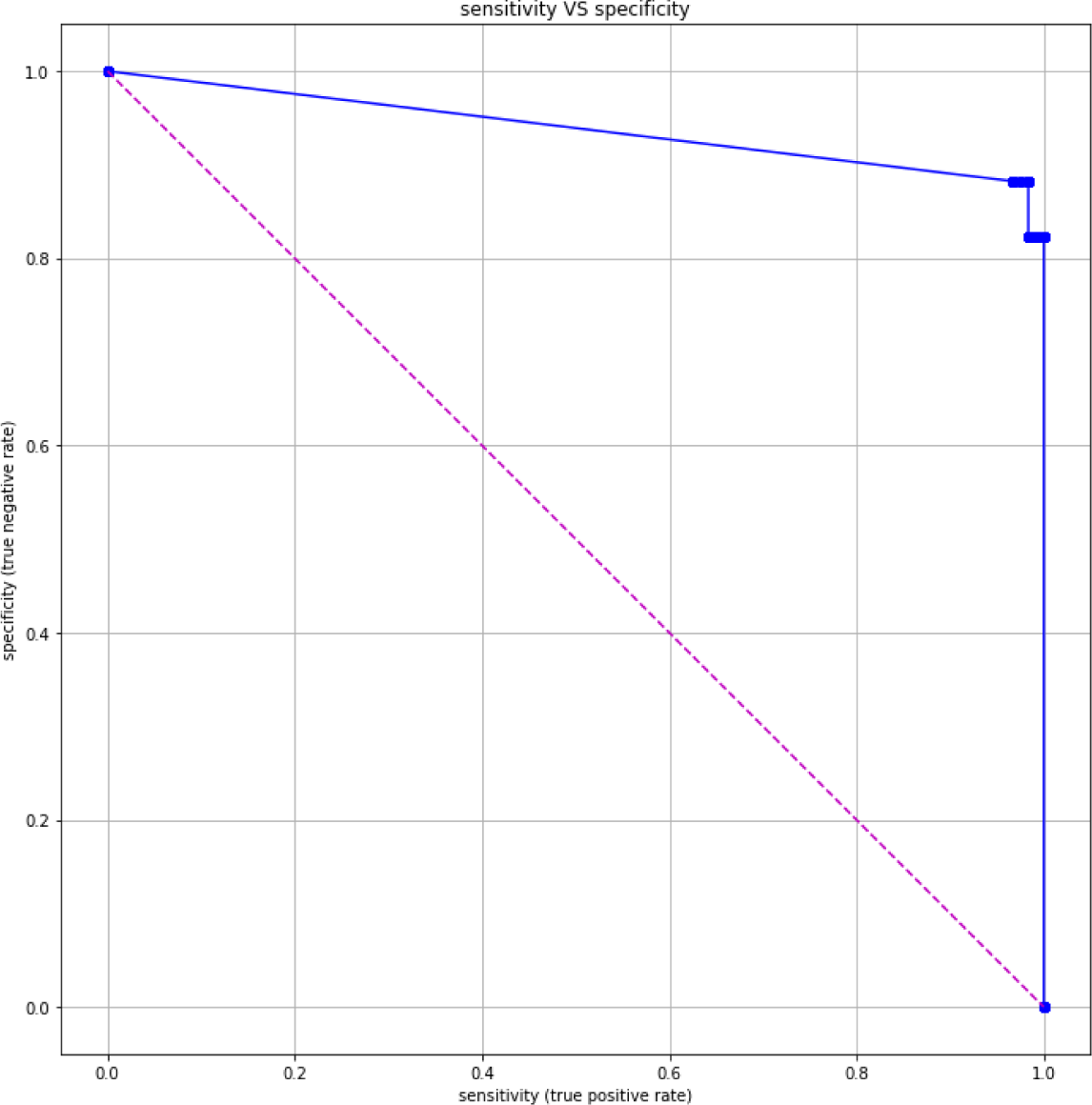
Sensitivity-specificity curve of the sepsis diagnosis at admission time.

## 4. Discussion

The identification of the variables involved in the diagnosis of sepsis (see Table 3) reveals that histone H2B plays an important role. Furthermore 3 of the 6 indicators used today in sepsis diagnosis are also identified by the algorithm: systolic blood pressure, procalcitonin levels and Breathing frequency. These results are consistent but would require more patients to be confirmed.

Regarding the predictive model performance, it exhibits encouraging BACC and AUC in section 3. Therefore, it should be emphasized that the number of available observations (the N) is low. This lack of samples in addition with the difference in the number of individuals per group (class imbalance: 17 controls against 122 cases) leads to think that the predictive models may not generalize as good as it would be desirable, behaving inappropriately with new patients. Therefore, a validation phase with new participants will be necessary to confirm the results obtained in this study.

## 5. Conclusions

From a database of 139 patients and 56 variables (demographic, treatment, etc.) a predictive model based on machine learning techniques to help diagnosing sepsis in patients admitted to the emergency room has been developed. Moreover, a set of relevant variables have been identified where 1 of the 4 variables of the HistShock® test appears as relevant, and the performance of the model is encouraging with a BACC of 97%. Nevertheless, these preliminary results should be confirmed with new patients.

## Data Availability

All data produced in the present study are available upon reasonable request to the authors.

## 6. Acknowledgements

We acknowledge INCLIVA and Hospital Clínico Universitario de Valencia (Valencia, Spain) to facilitate access to the dataset.

## Footnotes

1 Instituto Tecnológico de Informática, Valencia, Spain https://www.iti.es/

